# Treatment planning for photodynamic therapy of abscess cavities using patient-specific optical properties measured prior to illumination

**DOI:** 10.1101/2023.10.23.23297420

**Authors:** Zihao Li, Md Nafiz Hannan, Ashwani K. Sharma, Timothy M. Baran

## Abstract

**Background:** Photodynamic therapy (PDT) is an effective antimicrobial therapy that we used to treat human abscess cavities in a recently completed Phase 1 clinical trial. This trial included pre-PDT measurements of abscess optical properties, which affect the expected light dose to the abscess wall and eventual PDT response.

**Purpose:** The objective of this study was to simulate PDT treatment planning for the 13 subjects that received optical spectroscopy prior to clinical abscess PDT. Our goal was to determine the impact of these measured optical properties on our ability to achieve fluence rate targets in 95% of the abscess wall.

**Methods:** During a Phase 1 clinical trial, 13 subjects received diffuse reflectance spectroscopy prior to PDT in order to determine the optical properties of their abscess wall. Retrospective treatment plans seeking to achieve fluence rate targets in 95% of the abscess wall were evaluated for all subjects for 3 conditions: (1) at the laser power delivered clinically with assumed optical properties, (2) at the laser power delivered clinically with measured optical properties, and (3) with patient-specific treatment planning using these measured optical properties. Factors modified in treatment planning included delivered laser power and intra-cavity Intralipid (scatterer) concentration. The effects of laser fiber type were also simulated.

**Results:** Using a flat-cleaved laser fiber, the proportion of subjects that achieved 95% abscess wall coverage decreased significantly when incorporating measured optical properties for both the 4 mW/cm^2^ (92% vs. 38%, p=0.01) and 20 mW/cm^2^ (62% vs. 15%, p=0.04) fluence rate thresholds. However, when measured optical properties were incorporated into treatment planning, a fluence rate of 4 mW/cm^2^ was achieved in 95% of the abscess wall for all cases. In treatment planning, the optimal Intralipid concentration across subjects was found to be 0.14 ± 0.09% and the optimal laser power varied from that delivered clinically but with no clear trend (p=0.79). The required laser power to achieve 4 mW/cm^2^ in 95% of the abscess wall was significantly correlated with measured µ_a_ at the abscess wall (ρ=0.7, p=0.008), but not abscess surface area (ρ=0.2, p=0.53). When using spherical diffuser fibers as the illumination source, the optimal intralipid concentration decreased to 0.028 ± 0.026% (p=0.0005), and the required laser power decreased also (p=0.0002), compared to flat cleaved fibers. If the intra-cavity lipid emulsion (Intralipid) was replaced with a non-scattering fluid, all subjects could achieve the 4 mW/cm^2^ fluence rate threshold in 95% of the abscess wall using a spherical diffuser, while only 69% of subjects could reach the same criterion using a flat cleaved fiber.

**Conclusions:** The range of optical properties measured in human abscesses reduced coverage of the abscess wall at desirable fluence rates. Patient-specific treatment planning including these measured optical properties could bring the coverage back to desirable levels by altering the Intralipid concentration and delivered optical power. These results motivate a future Phase 2 clinical trial to directly compare the efficacy of patient-specific-treatment planning with fixed doses of Intralipid and light.

*Clinical Trial Registration:* The parent clinical trial from which these data were acquired is registered on ClinicalTrials.gov as “Safety and Feasibility Study of Methylene Blue Photodynamic Therapy to Sterilize Deep Tissue Abscess Cavities,” with ClinicalTrials.gov identifier NCT02240498.

## Introduction

An abscess consists of a fibrous pseudo-capsule surrounding a localized collection of bacteria, purulent fluid, and necrotic tissue^1^. Abscesses form as a result of the immune and inflammatory response to an acute infection, leading to nausea, pain, and high morbidity and mortality if untreated^2^. While the application of image-guided percutaneous abscess drainage has become the standard of care, response rate can be highly variable between subjects^3–5^. Additionally, there is concern that the antibiotics used alongside drainage will become less effective in the future, as antibiotic-resistant bacteria become increasingly common. We and others have directly observed the presence of multidrug-resistant bacteria in fluid aspirated from human abscesses^6–8^. It is therefore apparent that alternative treatment strategies are necessary, particularly for antibiotic-resistant strains.

Photodynamic therapy (PDT), which generates cytotoxic reactive oxygen species via light activation of drugs known as photosensitizers, may represent an ideal candidate treatment. PDT is a highly effective antimicrobial therapy that maintains efficacy against antibiotic resistant bacteria^9^, and can additionally improve susceptibility to antibiotics^10^. Based on encouraging pre- clinical results showing that PDT with methylene blue (MB) is efficacious against multiple bacteria typically found in abscesses^6,11,12^, we initiated a Phase 1 clinical trial investigating the safety and feasibility of MB-PDT at the time of abscess drainage (ClinicalTrials.gov identifier: NCT02240498). We found that MB-PDT was safe, with no study-related adverse events observed^8^. Additionally, preliminary analysis suggests that symptom resolution was faster and drainage catheter discharge was reduced in subjects receiving higher fluence. This clinical study used a uniform MB concentration of 1 mg/mL (0.1%) across subjects and the optical power delivered was based purely upon abscess geometry, rather than measured optical properties. During delivery of treatment light, each abscess was filled with a uniform 1% concentration of lipid emulsion in order to homogenize the light dose at the abscess wall through scattering. This scattering is critical to light delivery, as abscess morphology can be highly heterogeneous.

While we based the optical power delivered clinically on irradiance values that were efficacious *in vitro*^6,12^, the absorbed light dose is of course dependent upon the optical properties (absorption and scattering) of the target tissue. Multiple studies have shown that incorporation of patient-specific optical properties into PDT treatment planning results in improved coverage of the target region^13–15^. This is particularly true for the case of hollow cavities^16,17^, where the integrating sphere effect can dramatically increase fluence at the cavity boundary^18^. Lilge *et al*, for example, showed that this integrating sphere effect leads to a multiplicative factor in fluence rate, which is patient-specific and highly dependent on optical properties^19^. We have previously looked at simulated PDT of abscess cavities using our Monte Carlo simulation package^20,21^. We found that the ability to deliver an efficacious light dose, and the optical power required to do so, are highly dependent upon the optical properties at the abscess wall.

Whereas prior abscess treatment planning studies utilized assumed optical properties^20,21^, we have now measured the optical properties of 13 human abscesses immediately prior to PDT using a spatially-resolved diffuse reflectance spectroscopy system^22,23^. These measurements were performed before and after methylene blue administration, in order to capture bulk abscess wall optical properties and MB uptake. Using these pre-PDT optical property measurements and pre-procedure CT imaging, it is now possible to generate patient-specific treatment plans for subjects that received PDT of their abscess.

In this study, we describe the generation of patient-specific treatment plans based on pre-procedure CT imaging and intra-procedure measurement of optical properties. Specifically, we investigate the effects of measured optical properties on the delivered light dose, and determine whether patient-specific treatment planning can improve dose volume histograms in the presence of high abscess wall absorption. Further, we explore the impact of treatment fiber type and elimination of the intra-cavity lipid emulsion. We hypothesize that patient-specific treatment planning will increase the percentage of the abscess wall that achieves a target fluence rate, and that treatment planning will aid in optimization of MB and lipid emulsion concentrations for future studies.

## Methods

### Study participants and regulatory approval

Research subjects were part of a Phase 1 clinical trial examining the safety and efficacy of methylene blue (MB) mediated photodynamic therapy (PDT) delivered at the time of percutaneous abscess drainage. The full details and results of this clinical trial are described elsewhere^8^. As part of this Phase 1 trial, 13 subjects received diffuse reflectance spectroscopy prior to MB-PDT in order to determine the optical properties of their abscess, as described below. The present study focuses on the examination of treatment planning for these 13 subjects, including patient-specific optical properties and pre-procedure CT imaging.

Human data were collected as part of a Phase 1 clinical trial, which was approved by the Research Subjects Review Board at the University of Rochester Medical Center (IRB protocol number: STUDY00000488). This clinical trial was registered on ClinicalTrials.gov (ClinicalTrials.gov identifier: NCT02240498). All subjects provided written informed consent.

### Optical spectroscopy data

As part of the Phase 1 clinical trial, subjects first received standard of care image-guided percutaneous abscess drainage. Following this, an initial diffuse reflectance spectroscopy measurement of the abscess wall was made using a custom fiber-optic probe and diffuse reflectance spectroscopy system. These components, as well as the Monte Carlo lookup table approach for optical property recovery, were previously described by Bridger *et al*^24^. A full description of the clinical measurement protocol is provided elsewhere^22,23^. This pre-MB measurement represents the native optical properties of the abscess wall.

Following the initial measurement, MB was instilled into the cavity at a concentration of 1 mg/mL and allowed to incubate for 10 minutes. The cavity was then rinsed with sterile saline, and diffuse reflectance measurements were repeated to quantify abscess wall optical properties including the effects of MB absorption. Optical properties extracted from these post-MB measurements are the ones used below for the layer representing MB uptake by bacteria and diffusion into tissue.

Full details on abscess wall optical properties and MB uptake are described elsewhere by Hannan *et al*^22,23^. Here, we focus on the effects of these extracted optical properties on patient-specific treatment planning to optimize light delivery to individual abscess cavities.

### Pre-procedure imaging and segmentation

As part of their standard of care, all subjects received pre-procedure CT imaging no more than 7 days prior to image-guided percutaneous abscess drainage. These images were downloaded in a de-identified fashion from the picture archiving and communication system (PACS) at the University of Rochester Medical Center, and stored on a password-protected, encrypted workstation. Abscess location was confirmed on imaging by the study doctor (A.K.S.). Each set of CT images was then manually segmented by a member of the study team using Amira (Amira 3D v2022.1, ThermoFisher Scientific, Waltham, MA). This was done individually for each axial slice, with the abscess defined as a region of low density surrounded by a highly enhancing rim. Segmented abscesses were exported as DICOM files for inclusion in the treatment planning software described below.

### Treatment planning software

Measured human optical properties and corresponding segmented CT images were imported into our treatment planning software^20,21^, which is based upon our previously described Monte Carlo package^25^. This software represents patient anatomy with cuboid voxels, with each voxel being assigned corresponding optical properties. Here, segmented images were represented in two ways:

1) Divided into four regions: the abscess wall, surrounding tissue, inside the abscess, and a boundary layer. The abscess wall, consisting of a 200 µm thick layer immediately surrounding the abscess, was assigned optical properties corresponding to post-MB spectroscopy. The thickness of this layer was based upon reports showing similar diffusion of MB into various tissue types after application at the surface^26–29^. The surrounding tissue was set to pre-MB optical properties. This was meant to represent MB uptake by bacterial biofilms and the diffusion of MB into the abscess wall, rather than assuming homogeneous distribution of MB throughout the abscess wall and surrounding tissue.
2) Divided into three regions: a combined region representing the abscess wall and surrounding tissue, inside the abscess, and a boundary layer. This is the approach used in previous reports by Baran *et al*^20^ and Li *et al*^21^. In this case, optical properties of the abscess wall and surrounding tissue were both assigned the values measured in post-MB spectroscopy measurements.

The illumination sources simulated were physically accurate models of either the flat-cleaved optical fiber used clinically or a spherical diffusing fiber with an outside diameter of 2 mm. In both cases, the distal end of the fiber was positioned at the center of mass of the abscess cavity, and a fluence rate of 1 mW was delivered by simulation of 10^7^ photon packets. Output fluence rate maps were scaled linearly to simulate higher delivered optical power, with a maximum value of 10,000 mW. All simulations were performed using a Quadro RTX 6000 graphics processing unit (NVIDIA Corporation, Santa Clara, CA).

### Simulation conditions

For both flat-cleaved and spherical sources, we simulated three conditions:

1) Assumed treatment – absorption (µ_a_) and scattering (µ_s_) coefficients for the abscess wall were set to the assumed values of µ_a_=0.2 cm^-1^ and µ_s_=100 cm^-1^. The lipid emulsion concentration was fixed at 1%, and the delivered laser power was set to that delivered clinically for each subject. This case was meant to represent the dose we assumed was delivered clinically, which did not include patient-specific optical properties.
2) Delivered treatment with measured optical properties – µ_a_ and µ_s_ were set to the optical properties measured for each individual subject, while the lipid emulsion concentration and laser power were fixed at the values used clinically. This case represents a simulation of the dose delivered clinically.
3) Treatment planning with measured optical properties - µ_a_ and µ_s_ were set to the optical properties measured for each individual subject, and the lipid emulsion concentration and delivered laser power were optimized as described in the previous section. This case represents the treatment plan that would have been utilized if the treatment protocol were able to be modified on a patient to patient basis.

For the optimized treatment plan case, we simulated lipid emulsion concentrations ranging from 0-1%. The concentration used clinically was 1%, and we previously found that lower concentrations were optimal in a retrospective simulation study^21^. The assumed values of µ_a_=0.2 cm^-1^ and µ_s_=100 cm^-1^ were based upon values reported for tissues in the peritoneal cavity^30^. Scattering anisotropy was set to 0.7 inside the abscess, corresponding to the value for Intralipid at 665 nm^31^, and 0.9 in the abscess wall and surrounding tissue.

In all cases, the goal was to determine the delivered optical power (threshold optical power) and lipid emulsion concentration necessary to achieve a desired fluence rate of 4 mW/cm^2^ or 20 mW/cm^2^ in 95% of the abscess wall, while limiting the portion of the wall that receives a fluence rate of 400 mW/cm^2^ to less than 5%. The 4 mw/cm^2^ target is based upon an efficacious pre- clinical condition^6,12^, while the 20 mW/cm^2^ target is the target fluence rate used clinically^8^ to account for the more demanding *in vivo* scenario. The final treatment plan was determined by the lipid emulsion concentration that minimized the threshold optical power.

### Statistical analysis

For each of the simulation conditions described above, the following endpoints were calculated: (1) percentage of the abscess wall receiving at least 4 mW/cm^2^, (2) percentage of the abscess wall receiving at least 20 mW/cm^2^, (3) percentage of the abscess wall receiving at least 400 mW/cm^2^, and (4) delivered optical power required to achieve the threshold fluence rates of 4 and 20 mW/cm^2^. Comparison of these between treatment planning conditions was performed with the Wilcoxon test. Differences in the proportions of subjects that achieved the desired 95% abscess wall coverage at a given fluence rate were compared using Fisher’s exact test. Correlation between endpoints and abscess characteristics were calculated using the Spearman correlation coefficient. Statistical analysis was performed in GraphPad Prism (v9, GraphPad Software, Inc., Boston, MA) and MATLAB (R2022b, The Mathworks, Inc., Natick, MA).

## Results

### Representative case

A representative case is shown in Figure 1. A single slice of the segmented CT scan is shown in Figure 1a, and a rendering of the full segmentation is shown in Figure 1b. For this particular subject, optical properties at the treatment wavelength of 665 nm were found to be µ_a_ = 0.08 cm^-1^ and µ_s_’ = 4.8 cm^-1^ prior to MB administration, and µ_a_ = 49.4 cm^-1^ and µ_s_’ = 3.0 cm^-1^ following MB administration. For the 528 mW delivered clinically and the clinically deployed 1% Intralipid concentration, this resulted in 49.4% of the abscess wall achieving a fluence rate of 4 mW/cm^2^ with a flat-cleaved fiber. Simulations were then run for Intralipid concentrations ranging from 0-1% Intralipid, as described above. As shown in Figure 1c, the optimal Intralipid concentration was found to be 0.083%, with a corresponding optical power of 3914 mW. As shown in Figure 1d, this combination resulted in 95% of the abscess wall achieving the fluence rate target of 4 mW/cm^2^, while only 1.9% experienced a fluence rate of at least 400 mW/cm^2^.

**Figure 1.**
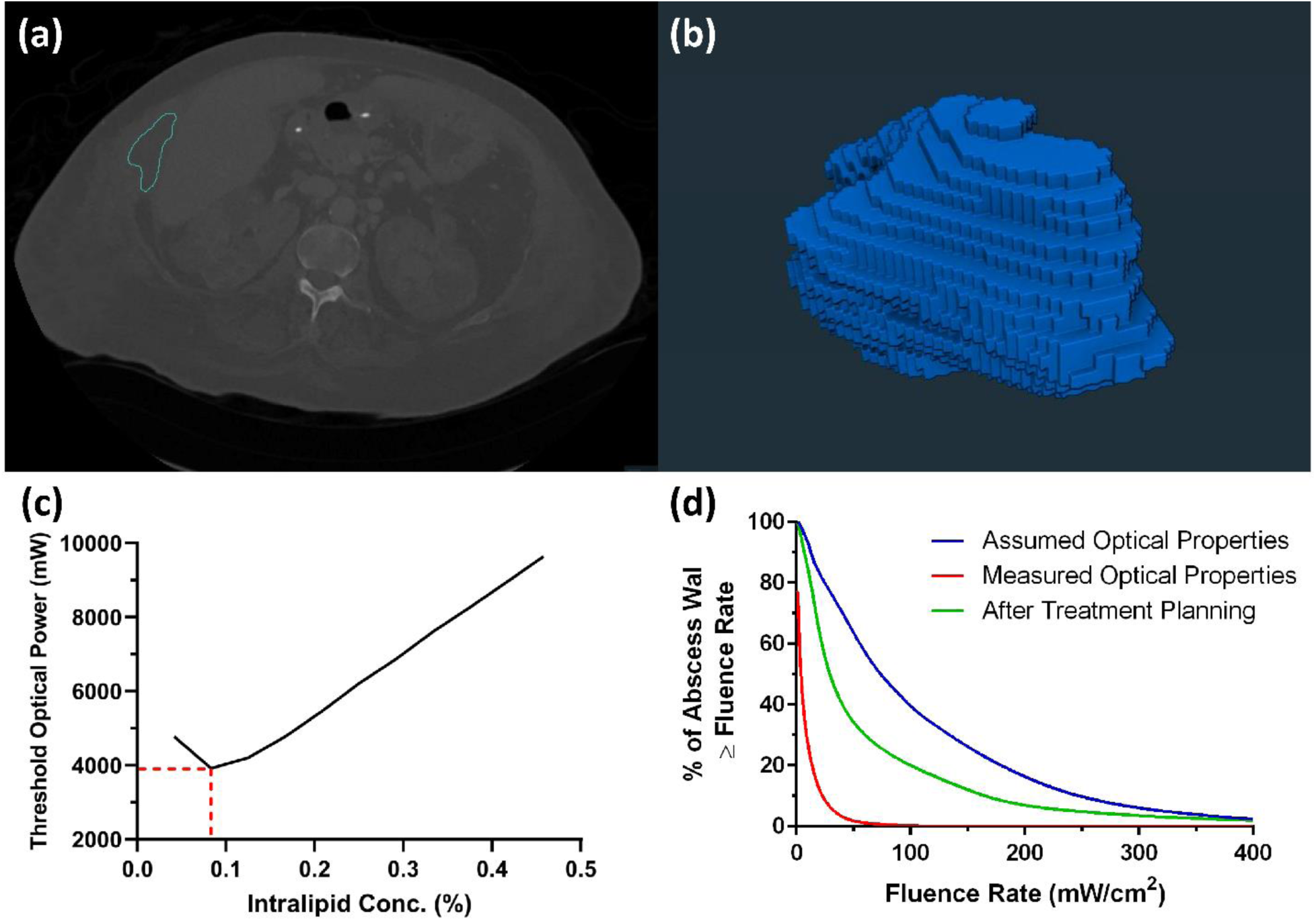
(a) Pre-procedure Computed Tomography (CT) image for a representative subject. The margin of the segmented abscess area is shown in light blue. (b) Segmented abscess rendered in three dimensions (3D) for the same individual. (c) Threshold optical power as a function of Intralipid concentration inside the abscess. The intersection of the red horizontal and vertical dashed lines indicates the optimal Intralipid concentration and corresponding optimal optical power for this subject after treatment planning. (d) Dose volume histogram representing coverage of the abscess wall as a function of fluence rate for the three simulated conditions.

### Treatment planning for flat-cleaved optical fibers

Across all 13 subjects, the target fluence rate of 4 mW/cm^2^ was reached in 95% of the abscess wall using assumed optical properties (µ_a_=0.2 cm^-1^, µ_s_’=10 cm^-1^) and the clinically delivered laser powers in all but one subject (Figure 2a,b). When measured optical properties were simulated with 1% Intralipid and the laser powers delivered clinically, abscess wall coverage at 4 mW/cm^2^ decreased significantly (p=0.005, Figure 2a,b), with only 5 subjects attaining the desired 95% coverage. However, when measured optical properties were incorporated into treatment planning (TP), a fluence rate of 4 mW/cm^2^ was achieved in 95% of the abscess wall for all cases. Further, treatment planning decreased the portion of the abscess wall receiving fluence rates exceeding 400 mW/cm^2^ (Figure 2e), and reduced the number of subjects expected to experience this high fluence rate in greater than 5% of the abscess wall.

**Figure 2.**
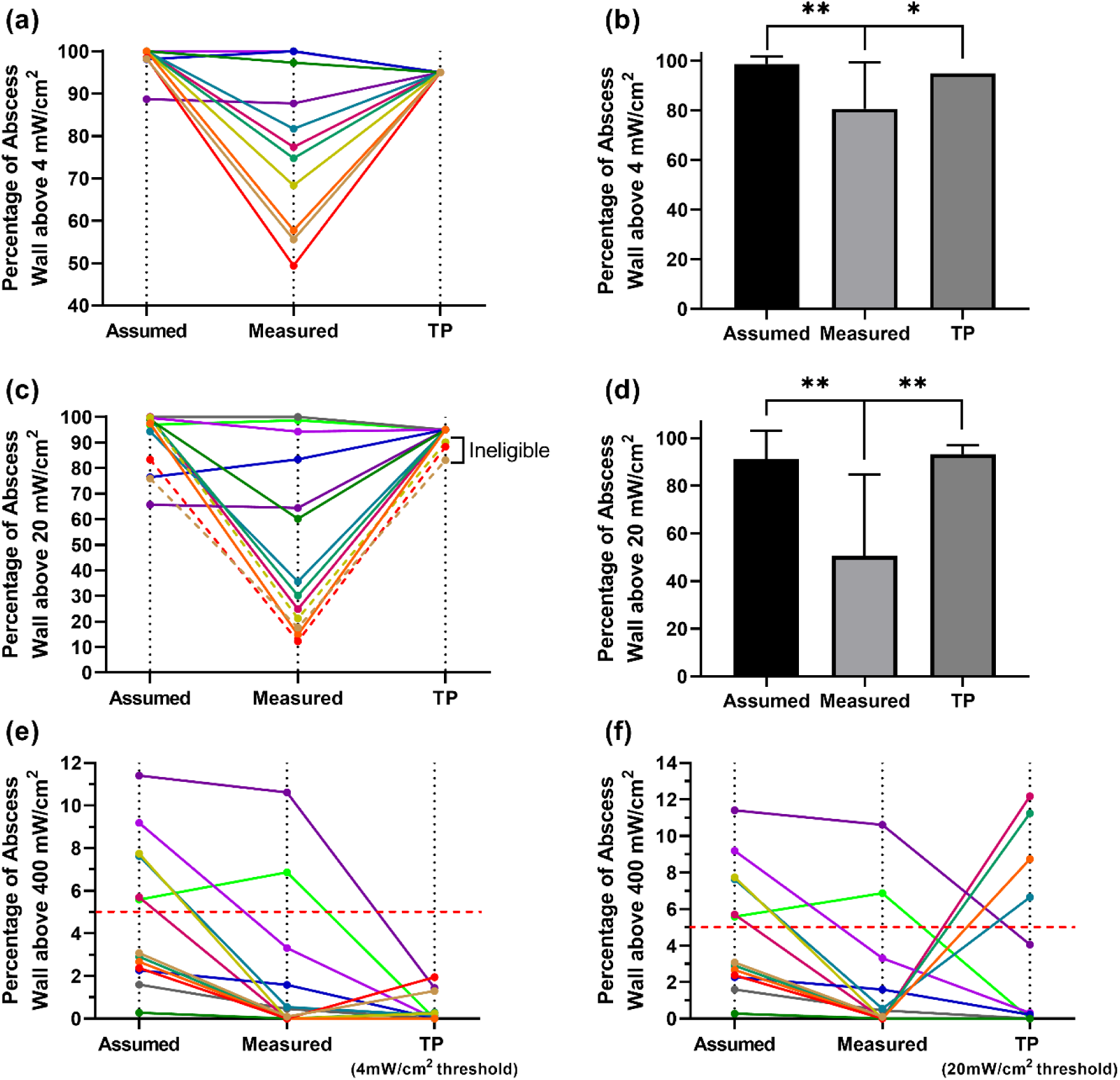
Percentage of the abscess wall that received a fluence rate above (a) 4 mW/cm^2^ or (c) 20 mW/cm^2^ under assumed, measured, and treatment planning cases for all 13 subjects. Each color represents an individual subject. Corresponding means and standard deviations across subjects are included for the (c) 4 mW/cm^2^ and (d) 20 mW/cm^2^ cases. Percentage of the abscess wall that received a fluence rate above 400 mW/cm^2^ for these three cases at thresholds of (e) 4 mW/cm^2^ and (f) 20 mW/cm^2^. * p<0.05 ** p<0.01

This treatment planning improvement was enabled by a reduction in Intralipid concentration relative to the 1% concentration deployed clinically and changes in delivered laser optical power. The mean optimal Intralipid concentration across subjects, determined using the method shown in Figure 1c, was found to be 0.14 ± 0.09% with a range of 0-0.4%. As seen in Figure 3, the optimal laser power was typically different from that delivered clinically, with some cases requiring greater laser power than was used clinically and some requiring less. There was a slight increase in threshold optical power for treatment planning relative to clinical values (Figure 3), although this difference was not significant (p=0.79).

**Figure 3.**
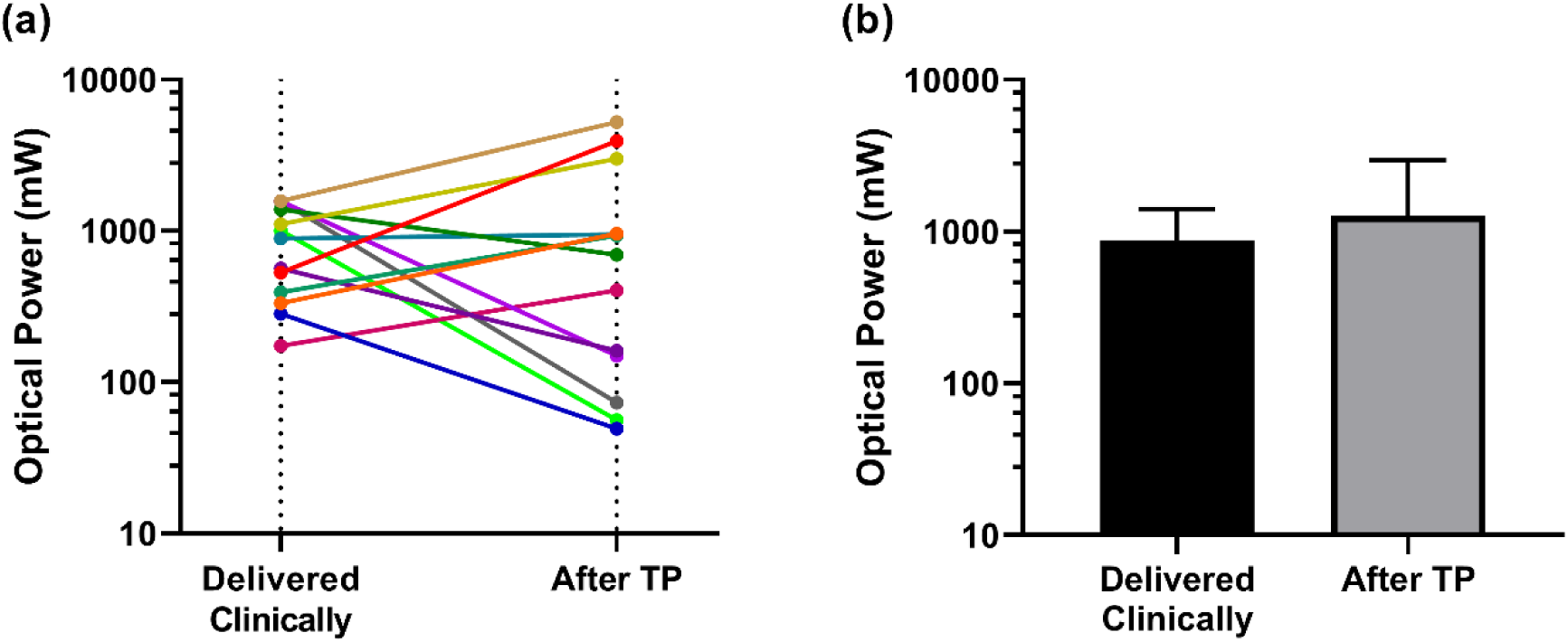
(a) Comparison of the optical power that was delivered clinically and optimal optical power determined by treatment planning for all 13 subjects. Each color represents an individual subject. (b) Corresponding means and standard deviations across subjects.

We found that the required laser power to achieve 4 mW/cm^2^ in 95% of the abscess wall was significantly correlated with measured µ_a_ at the abscess wall (ρ=0.7, p=0.008; Figure 4a). However, this required power was not significantly correlated with abscess surface area (ρ=0.2, p=0.53; Figure 4b). This indicates that the effects of abscess wall optical properties on required illumination power are more pronounced than the effects of abscess size.

**Figure 4.**
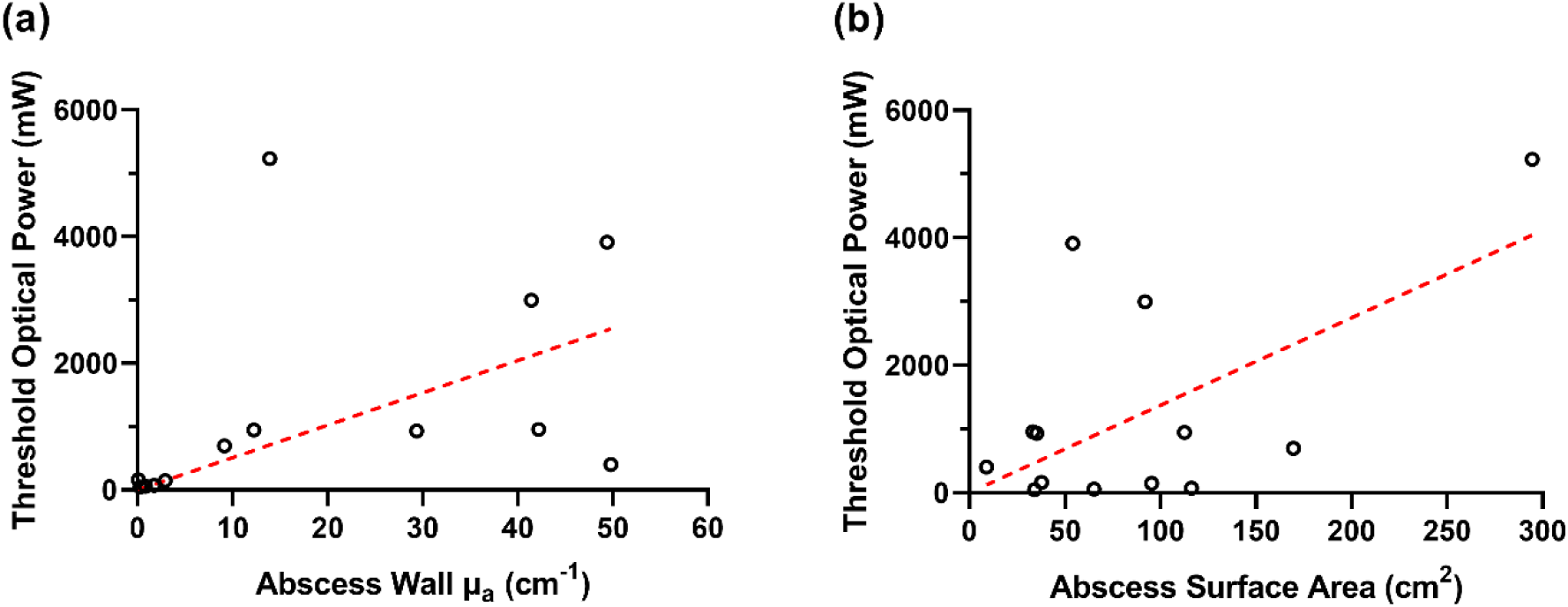
Relationship between optical power required to achieve a fluence rate of 4 mW/cm^2^ in 95% of the abscess wall after treatment planning and (a) abscess wall µ_a_, or (b) abscess surface area, using flat cleaved fiber.

Similar trends were seen for the 20 mW/cm^2^ fluence rate target. As with the 4 mW/cm^2^ condition, there was a significant decrease in the number of subjects attaining 95% coverage when applying measured optical properties and using the laser powers delivered clinically (p=0.008, Figure 2c,d). However, the number of cases where 95% coverage was not attainable for both assumed and measured optical properties increased compared to the 4 mW/cm^2^ case (Figure 2c). Additionally, there were three subjects for which 95% coverage at 20 mW/cm^2^ could not be achieved with treatment planning.

### Treatment planning for spherical diffusers

In addition to performing treatment planning for the flat-cleaved optical fibers used to deliver laser illumination clinically, we also examined the use of spherical diffusers for light delivery. Similar trends were observed, with lower coverage of the abscess wall at a specific fluence rate threshold when measured optical properties were simulated, relative to assumed optical properties. Additionally, after treatment planning the 4 mW/cm^2^ and 20 mW/cm^2^ fluence rate thresholds were achievable in 95% of the abscess wall for all cases. However, the spherical diffuser case required lower optical power and had a lower optimal Intralipid concentration compared to the flat-cleaved fiber results (Figure 5). The optimal Intralipid concentration decreased significantly from 0.14 ± 0.09% to 0.028 ± 0.026% (p=0.0005, Figure 5a,c), and the required laser power decreased significantly from the flat-cleaved case (p=0.0002, Figure 5b,d).

**Figure 5.**
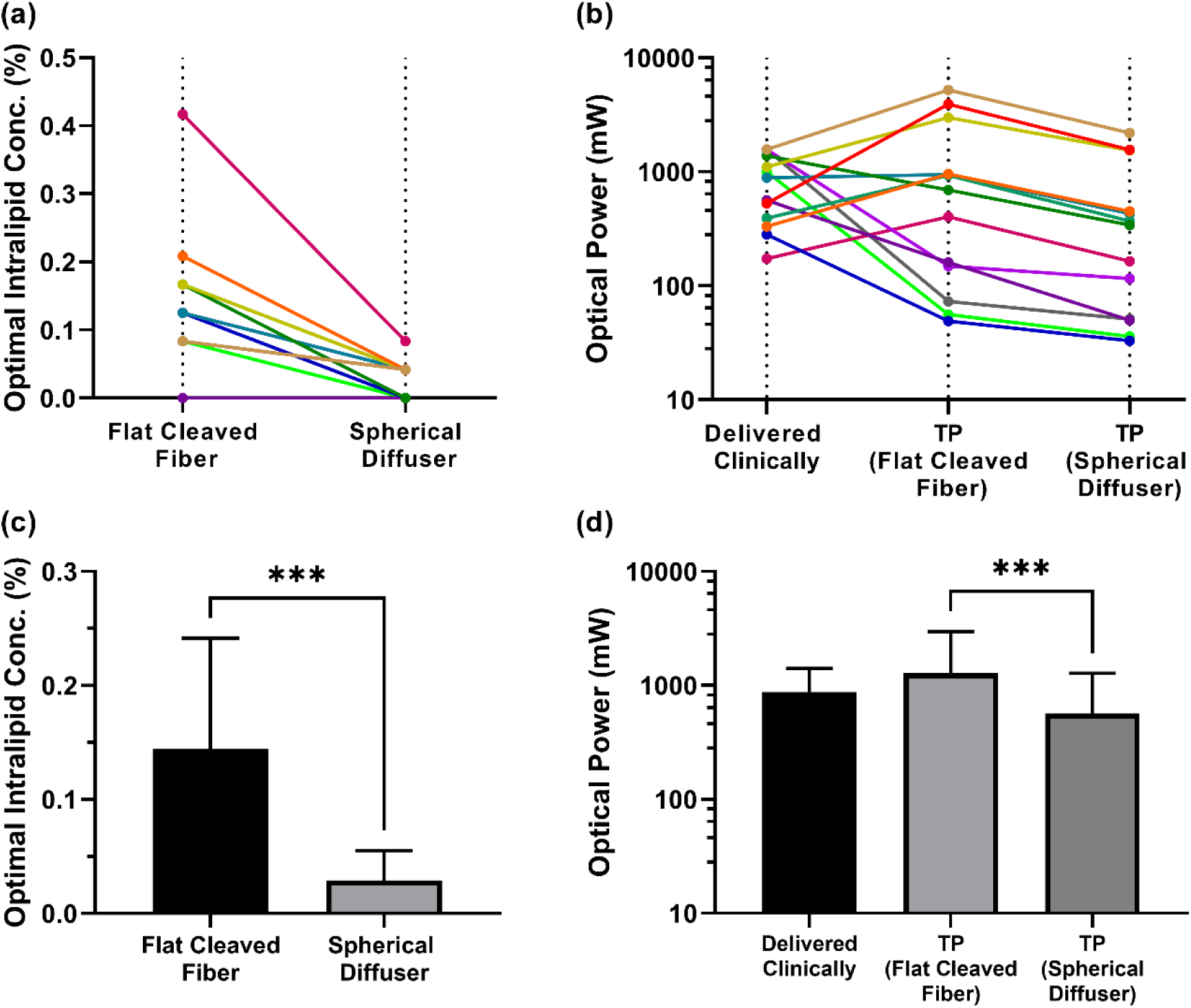
Comparison between flat-cleaved and spherical diffuser fibers for (a) optimal Intralipid concentration and (b) required optical power after treatment planning. Corresponding means and standard deviations are included for (c) optimal Intralipid concentration and (d) required optical power. *** p<0.001

### Effects of eliminating intra-cavity lipid emulsion

While our clinical study included 1% Intralipid within the abscess cavity during illumination and we have found that a non-zero Intralipid concentration is optimal for both flat-cleaved and spherical diffuser fibers, we also simulated the case of 0% Intralipid within the cavity. This represents the case where a non-scattering index-matching fluid is infused into the cavity.

Across all 13 subjects, 69% of subjects could reach the 4 mW/cm^2^ fluence rate threshold in 95% of the abscess wall after treatment planning using a flat cleaved fiber, while all subjects could achieve the same criterion using a spherical diffuser. As expected, these results were more favorable for the spherical diffuser (Figure 6), as the flat-cleaved fiber requires intra-cavity scattering to reduce the forward-peaked nature of light emitted from the fiber. For flat-cleaved fibers, a greater laser power was typically required to achieve the fluence rate target, relative to conditions including intra-cavity scattering. For spherical diffuser fibers, the required optical power was decreased relative to conditions including scattering (p=0.004).

**Figure 6.**
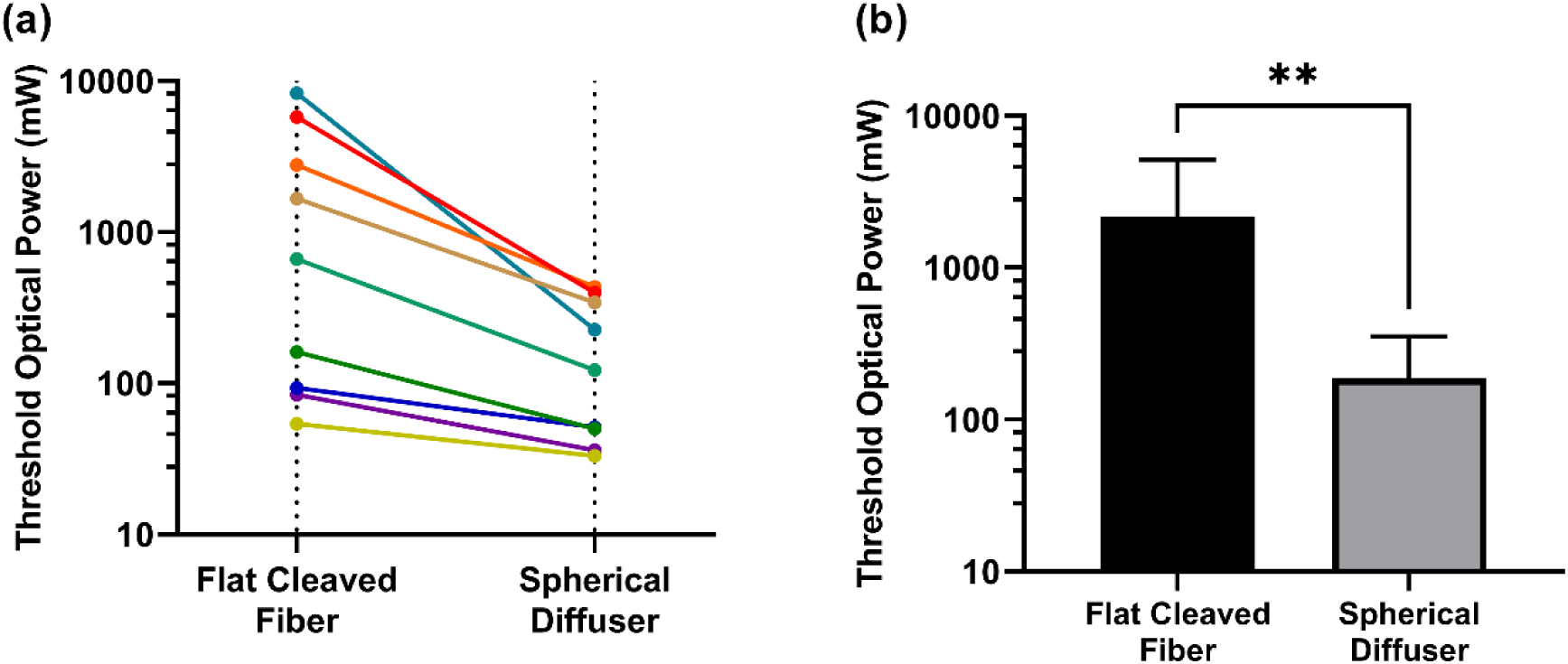
(a) Comparison of threshold optical power after treatment planning between flat cleaved fibers and spherical diffuser fibers for the case of 0% Intralipid concentration with (b) corresponding means and standard deviations across subjects. ** p<0.01

### Assumption of post-MB optical properties

All prior results reported here simulated the effects of MB penetration into the abscess wall by assigning optical properties measured after MB instillation and aspiration to a 200 µm thick layer at the abscess wall. Surrounding tissue optical properties were set to those measured prior to the administration of MB. This was meant to simulate the relatively shallow penetration of MB into tissue observed in prior studies^26–29^. To examine the effects of this assumption, we also simulated an artificial condition where optical properties measured post-MB were assigned to the abscess wall and surrounding tissue. As would be expected, abscess wall coverage decreased markedly relative to the 200 µm layer case. Further, 95% coverage of the abscess wall could only be achieved in 31% of cases for the 4 mW/cm^2^ threshold as opposed to 100% in the 200 µm layer case.

## Discussion

In this study, we found that simulation of PDT of deep tissue abscesses with optical properties measured in treated human subjects reduced the proportion of the abscess wall attaining a given fluence rate target, relative to assumed optical properties. However, by applying patient-specific treatment planning using these measured optical properties and modifying the Intralipid concentration present within the abscess during PDT and the delivered laser power, the desired 95% coverage of the abscess wall could be recovered. This was true for both the clinically utilized flat-cleaved optical fibers and spherical diffuser fibers, with spherical fibers potentially allowing for lower laser power and reduced Intralipid concentration. These results motivate the prospective application of patient-specific treatment planning in future clinical trials of PDT for treatment of abscesses, and highlight the importance of optical property measurements prior to PDT.

We have previously shown that patient-specific treatment planning incorporating knowledge of optical properties improved eligibility for abscess PDT in a retrospective population using assumed ranges of tissue absorption and scattering^21^. Here, we demonstrated similar improvements using measured optical properties from subjects that received PDT at our institution. Other groups have also investigated dosimetry and treatment planning for PDT in hollow cavities, including for the bladder^19^, oropharynx/nasopharynx^17,32–34^, and pleural cavity^35,36^. Of note, many of these prior studies did not incorporate measured optical properties for individual patients, but instead either used assumed values or required insertion of separate fibers for dosimetry. This study therefore represents a step forward in patient-specific treatment planning for PDT of hollow cavities, including the effects of highly variable abscess wall optical properties and MB uptake.

While flat-cleaved optical fibers were used in the clinical setting for the subjects described here, we also simulated spherical diffuser fibers. We found that spherical diffusers allowed for lower laser power and lower Intralipid concentration to achieve the same coverage as flat-cleaved fibers. However, the main disadvantage of spherical diffusers is their reduced optical power threshold compared to flat-cleaved fibers. For example, the Medlight spherical diffuser (SD200, Medlight SA, Ecublens, Switzerland) can only handle an input optical power of 3 W in water or 1 W in air. For the abscesses treated here, the optical power delivered clinically was as high as 1571 mW. Anticipating treatment of larger abscesses in future studies, we are planning to utilize a laser system with output optical power as high as 10 W. These Medlight fibers would therefore be insufficient. However, alternative spherical diffusers, such as the Luminous Spherical Diffuser (Schott AG, Mainz, Germany), are rated for optical powers as high as 20 W. In addition to their higher power transmission capabilities, there is also the possibility to use flat-cleaved fibers for quantitative optical spectroscopy^37^. In this way, the additional fiber optic probe currently required could be eliminated in favor of an approach that uses the treatment fiber for both low power optical property measurement and delivery of high power treatment light. Validation of this approach is currently ongoing.

Based upon the findings reported here, the lipid emulsion concentration can clearly be lowered in future studies. The 1% lipid emulsion concentration used in the clinical study was based upon an apparent improvement in fluence rate distribution for higher MB concentrations at higher lipid emulsion concentration^20^. However, this finding has not been replicated here or in a prior simulation study^21^. The optimal lipid concentration was found to be 0.14 ± 0.09% for all subjects examined here, and 0-0.25% in a prior simulation study^21^. As other investigators have also found that lower intra-cavity lipid concentrations are most efficient for PDT of hollow cavities such as the bladder^19^, this concentration will likely be reduced in future clinical studies. Similarly, the MB concentration used clinically was likely too high, as evidenced by the large values of absorption at 665 nm. Further justification for use of lower MB concentrations is discussed in detail by Hannan *et al*^23^.

The exact effects of MB concentration are also dependent upon the penetration of MB into the target region during the drug-light interval. As we have shown here, simulation of a thin layer representing MB penetration results in a much larger proportion of subjects being eligible for PDT. However, the penetration depth assumed here is based upon reports in other tissue types for MB^26–29^, rather than those determined directly in abscesses. This motivates direct measurement of MB depth penetration into abscess tissue. This would be difficult or impossible in human subjects, as any damage to the abscess wall could result in abscess rupture and sepsis. However, animal models of abscess formation do exist in rabbits^38^ and sheep^39^. These model systems could be used to quantify MB depth penetration, similarly to what has been done for other photosensitizers in various sites^40–42^.

Some limitations are acknowledged in the execution of this study. These results are derived from a relatively small sample (n=13), so there is potential for selection bias. Since optical properties were only measured at a single location, we also make assumptions on homogeneity of optical properties. Tissue optical properties can be heterogeneous within a given region, and this heterogeneity can result in changes in absorbed light dose^43^. As our simulation framework can incorporate voxel-specific optical properties, future studies will focus on methods for mapping of heterogeneous optical properties and their effects on treatment planning. In this study, we also examined the effects of MB depth penetration on treatment planning. As described above, we do not have data on the penetration of MB into abscess tissue over the 10 minute drug-light interval employed in our clinical study. Inaccuracy in MB penetration depth could therefore affect the treatment planning results reported here. Finally, abscess morphology was based upon pre-procedure CT imaging. For the subjects described here, the interval between pre-procedure imaging and treatment varied from 0-7 days. It is therefore possible that abscess shape was slightly different at the time of PDT. This motivates utilization of CT imaging immediately prior to PDT, in order to capture accurate abscess morphology. This could be facilitated by real-time abscess segmentation, which has been demonstrated for orbital abscesses^44^.

## Conclusions

We determined that the ranges of optical properties measured in human abscesses result in decreased coverage of the abscess wall at a desired fluence rate for PDT. However, inclusion of these measured optical properties into patient-specific treatment planning returned delivered fluence rates to the desired levels by modification of intra-cavity Intralipid concentration and delivered laser power.

These findings motivate the performance of patient-specific treatment planning prospectively in future patients receiving PDT. It is anticipated that this will result in improved coverage of the abscess wall at an efficacious fluence rate, which will improve response to PDT and reduce negative clinical outcomes. In order to test this, we are currently planning a Phase 2 clinical trial that will directly compare PDT efficacy between a fixed light dose and patient-specific treatment planning.

## Data Availability

All data produced in the present study are available upon reasonable request to the authors

## Acknowledgments

The authors would like to thank Dr. Joan Adamo for regulatory support. This work was funded by grant EB029921 from the National Institutes of Health.

